# Wearable and Interview-based Assessment of Psychological Risk in Alzheimer’s Caregivers: Machine Learning vs. Large Language Models

**DOI:** 10.64898/2026.05.24.26353993

**Authors:** Junen Xiao, Zihan Zhao, Zachary D. King, Maryam Khalid, Sara Davies, Khadija Zanna, Daniel L. Argueta, Kelly N. Brice, E. Lydia Wu-Chung, Vincent D. Lai, Jensine Paoletti-Hatcher, Bryan T. Denny, Samantha Henry, Paul E. Schulz, Christopher P. Fagundes, Akane Sano

**Affiliations:** Department of Electrical and Computer Engineering, Rice University, Houston, TX, USA; Department of Computer Science, Rice University, Houston, TX, USA; Department of Psychological Sciences; Institute of Health Resilience and Innovation, Rice University, Houston, TX, USA; Department of Psychology, University of Pittsburgh, Pittsburgh, PA, USA; Department of Psychology, Portland State University, Portland, OR, USA; Section of Neuropsychology, Baylor College of Medicine, Houston, TX, USA; Department of Neurology, McGovern Medical School, UTHealth Houston, Houston, TX, USA

## Abstract

Spousal caregivers of individuals with Alzheimer’s disease and related dementias frequently experience elevated perceived stress, caregiver burden, and loneliness, which are associated with adverse health outcomes. Early identification is therefore critical for timely intervention. Existing approaches commonly rely on wearable sensor data and standardized psychological questionnaires, while recent multimodal methods aim to improve prediction by integrating behavioral and linguistic information.

In this study, we explored three modality configurations, wearable-derived features, interview-based text, and their combination, to classify caregiver psychological risk using the Perceived Stress Scale (PSS), Zarit Burden Interview, and UCLA Loneliness Scale. We compared traditional machine learning models and large language models (LLMs) (Gemini 2.0, Llama 4, and GPT-4o) under psychometrician-centered and caregiver-centered prompting strategies.

Traditional machine learning models performed better under multimodal settings, while LLMs achieved stronger performance with Interview-Only input. We further demonstrate that PSS was the most predictable construct and prompting strategies substantially influenced LLM performance.

**Author summary:** People caring for spouses with Alzheimer’s disease and related dementias often experience high levels of stress, caregiver burden, and loneliness, all linked to adverse psychological and physical health outcomes. Early identification of caregivers at heightened psychological risk is essential for timely support. We evaluated three data modalities, wearable-derived features, interview-based text, and their combination, to classify caregiver risk using the Perceived Stress Scale, Zarit Burden Interview, and UCLA Loneliness Scale. Traditional machine learning models and large language models (LLMs) (Gemini 2.0, Llama 4, GPT-4o) were compared under multiple prompting strategies.

Our findings showed that traditional machine learning approaches performed best when combining wearable-derived behavioral features with interview-derived linguistic features, while LLMs were more effective for analyzing interview-based text. PSS was the most predictable construct, while caregiver burden and loneliness were more difficult to detect. Prompting choices significantly influenced LLM performance, and Gemini 2.0 showed the most stable overall results. These findings highlight the importance of aligning model choice with data modality when developing digital health tools for caregiver risk identification.

## 1 Introduction

Caring for a spouse with Alzheimer’s Disease and Alzheimer’s Disease Related Dementias (AD/ADRD) constitutes a major source of psychological distress [1]. This chronic caregiving distress manifests across multiple dimensions, including perceived stress [2], caregiver burden [3], and loneliness [4], which capture distinct but interrelated aspects of the caregiving experience.

These psychological challenges are not only subjectively taxing but also associated with physiological responses, including increased pro-inflammatory activity and systemic inflammation [5, 6], which can lead to adverse health outcomes [7]. These effects often develop gradually and may not be immediately apparent in daily life [8] but their resilience and vulnerability vary substantially across individuals [9]. Therefore, timely and personalized assessment is essential for identifying caregivers at elevated risk and enabling targeted support.

Caregiver well-being is typically assessed using validated self-report instruments such as the Perceived Stress Scale (PSS) [10], Zarit Burden Interview (ZBI) [11], and UCLA Loneliness Scale (UCLALS) [12]. These instruments provide reliable measures of psychological states but are usually administered infrequently due to participant burden [13]. As a result, they may miss meaningful fluctuations in caregivers’ daily experiences. Moreover, the multidimensional nature of caregiver distress complicates assessment [14]: perceived stress reflects the appraisal of caregiving demands relative to coping resources [15], caregiver burden captures the subjective weight of caregiving responsibilities [16], and loneliness indexes perceived deficits in caregivers’ social connection [17]. These differences highlight the need for scalable, low-burden approaches capable of capturing both short-term variation and longer-term patterns in caregiver well-being.

Advances in digital health technologies offer new opportunities for continuous, multimodal assessment of psychological states. Wearable devices provide passive, high-frequency physiological and behavioral data [18], such as heart rate, sleep patterns, and circadian or other rhythms [19], which have been linked to stress and other mental health outcomes [20, 21]. Because perceived stress has well-documented physiological correlates—including changes in heart rate variability [22, 23] and sleep quality [24], wearable-derived features may be particularly informative for stress-related constructs. In contrast, loneliness primarily reflects subjective perceptions of social disconnection [17], and caregiver burden reflects cumulative task-related strain [25, 26], which may be less directly captured by physiological signals.

Text-based data provide a complementary window into caregivers’ emotional and cognitive states. NLP methods applied to written or spoken language can capture affective tone, cognitive patterns, and contextual information [27]. Traditional representations such as TF-IDF [28] and Word2Vec embeddings [29] have been used to predict stress [30], caregiver burden [31], and loneliness [32]. Stress is often explicitly expressed in daily language [33, 34], whereas loneliness and burden may involve more complex or context-dependent cues [35, 36], making them harder to detect from text alone.

Large language models (LLMs) also offer new capabilities for interpreting complex linguistic and contextual signals relevant to mental health [37]. Recent studies have explored prompting strategies, including zero-shot, few-shot, and emotion-aware prompting, to guide LLMs in predicting psychological states from social media text [38, 39]. Role-based prompting, in which LLMs adopt specific perspectives (e.g., counselor or clinician), has also been shown to influence reasoning patterns and improve task performance [40, 41]. However, most prior work relies on unstructured social media data, which may lack the coherence and contextual richness of structured interviews [42]. Little is known about how LLMs perform when applied to interview transcripts or when provided with multimodal inputs that combine text with wearable-derived features.

Motivated by these gaps, we conduct a systematic comparison of traditional machine learning models and LLMs for classifying AD/ADRD spousal caregivers into high-versus low-risk groups on the PSS, ZBI, and UCLALS scales. We evaluate multiple modality configurations, including wearable-only, interview-only, and combined text-plus-wearable inputs, and design caregiver-centered and psychometrician-centered prompting strategies that guide LLMs (Gemini 2.0, Llama 4, and GPT-4o) to perform the same classification task. Finally, we perform statistical analyses to compare model performance across methods and modalities, providing a comprehensive assessment of how different computational approaches capture distinct dimensions of caregiver well-being.

## 2 Methods

### 2.1 Study Design

A total of 43 caregivers of patients with Alzheimer’s disease were initially enrolled in the study. Among them, 41 participants completed a 28-question interview. To enable reliable computation of rhythm (< 24 hours, 24 hours, > 24 hours) features, we retained only participants with at least five valid days of complete 24-hour recordings, resulting in a final sample of 32 participants (28 female and 4 male). The study is approved under Rice University Institutional Review Board (IRB).

Inclusion criteria for AD spousal caregivers are: 1) self-identified as the principal person taking care of their spouse with dementia. 2) devoting at least 4 hours daily to the care of the spouse for at least the last 3 months. 3) the AD spousal caregiver & AD patient must have been married (or self-defined as long-term committed partner) for at least 3 years.

Participants wore Fitbit devices during a 7-day study block, which continuously recorded minute-level heart rate and step count data throughout the day (up to 1,440 samples per day), while sleep status was also recorded at minute-level resolution but only during detected sleep periods.

At the end of the study, participants completed a 30-minute semi-structured interview covering four thematic domains: 1) caregiving experiences, 2) appraisal of stressful events, 3) caregiving-related concerns, and 4) stress and coping strategies. Participants also completed the standardized surveys (PSS-10 [10], UCLALS Version [12], and ZBI-13 [11]), which served as ground-truth labels for perceived stress, loneliness, and caregiver burden, respectively.

### 2.2 Interview Data Processing

Transcripts were generated offline from the interview audio using OpenAI Whisper [43]. Coders reviewed the text, segmented the interviews into meaningful units, anonymized identifiable information, and cleaned the transcripts by removing filler words and other non-informative content. The final dataset comprises structured question–response transcripts spanning 28 predefined interview questions, systematically organized into thematic topics for downstream analysis.

### 2.3 Wearable Feature Extraction

We extracted sleep, daily, and rhythm features from the wearable data of each participant. Fig. 1 shows the feature extraction process.

**Fig 1.**
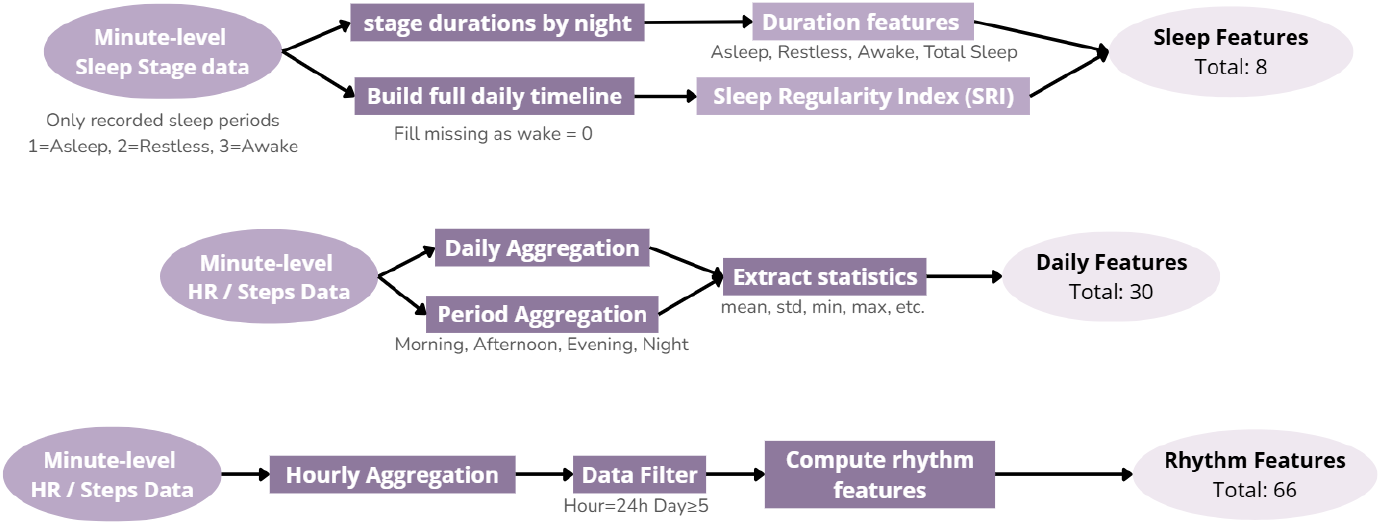
Wearable feature extraction pipeline

Sleep features mainly consist of the duration in different stages of sleep, including asleep, restless, and awake states. The Sleep Regularity Index (SRI) [44] was also calculated based on the whole-day sleep states at the minute-level resolution. The higher SRI indicates greater regularity in sleep patterns.

Daily features were derived from minute-level heart rate and step data, distress-related physiological responses, and daily activity patterns. Heart rate features included summary statistics and the proportion of minutes with elevated heart rate (HR>100 bpm). This is because elevated heart rate may indicate periods of heightened distress or physiological activation [45]. Activity features included step statistics, active and sedentary time proportions, and step entropy. Features were computed separately for different times of day (morning, afternoon, evening, and night).

Rhythm features were derived from hourly heart rate and step data to capture ultradian (< 24 h), circadian (24 h), and infradian (> 24 h) activity patterns. Circadian rhythms are strongly linked to health and well-being outcomes [21], while ultradian cycles (e.g., sleep-stage oscillations, cortisol pulses) and infradian cycles (e.g., menstrual and seasonal rhythms) also influence mood, arousal regulation, and distress sensitivity [46, 47].

M10 and L5 quantify the 10 most active and 5 least active hours per day [48], reflecting primarily circadian day–night structure with embedded ultradian variation. From these, rest–activity relative amplitude (RA) indexes circadian rhythm strength; lower or irregular RA indicates disrupted circadian and ultradian organization [48, 49]. To capture slower, multi-day changes, deviation-from-template features summarize how each day diverges from a 24-hour activity template computed over 7–28-day windows, representing infradian rhythm stability [49]. Finally, cosinor models with 8–24 h periods estimate MESOR (Midline Estimating Statistic of Rhythm), amplitude, and acrophase [50, 51]; 8–16 h fits capture ultradian cycles, while 24 h fits capture circadian rhythmicity. Together, these features quantify multi-timescale regularity in daily behavior.

In total, 104 features were extracted, including 8 sleep features, 30 daily features, and 66 rhythm features.

### 2.4 Baselines

We adopted two baseline methods for comparison with the proposed approaches. The majority-class baseline assigns all instances to the most frequent class in the training data, while the probabilistic random baseline generates predictions by sampling from the class distribution. Performance was evaluated by comparing the predicted binary labels with the ground truth labels.

### 2.5 Machine Learning Models

Given the small sample size and risk of overfitting in deep learning models, we employed traditional machine learning classifiers: Support Vector Machines (SVM), XGBoost (XGB), Random Forests (RF), and K-Nearest Neighbors (KNN). Models were trained to classify participants into high-vs. low-risk groups for PSS, ZBI, and UCLALS under different modality configurations.

We used Leave-One-Out Cross-Validation (LOOCV) due to the limited sample size (n = 32). In each fold, one participant served as the test case while the remaining 31 participants were used for training. Predictions across all folds were aggregated to compute evaluation metrics (S1 Fig).

Feature selection for wearable data was performed within each training fold. Low-variance features were removed, followed by correlation filtering and Random Forest–based importance ranking. The top five features were retained for model training. Text representations were generated using TF-IDF [28]. Feature extraction was performed within each training fold, and the learned transformations were applied to the held-out test sample without refitting. Multimodal features were constructed by concatenating wearable and text features within each fold.

We conducted feature contribution analyses using SHAP (SHapley Additive exPlanations) values for XGBoost (wearable features) and linear SVM coefficients for TF-IDF features (text). For LLM experiments, we selected a fixed set of five wearable features per scale based on selection frequency and importance (see S1 Table).

### 2.6 Prompt Engineering Strategies

We evaluated LLM-based classification using interview text, wearable-derived textual descriptions, or both. Wearable features were converted into natural-language statements containing feature names, explanations, and numeric values. For text-based modeling, we also tested prompts containing the top 20 TF-IDF features.

Because LLM outputs can vary depending on the instructed perspective [52], we designed two role-based prompting strategies: (1) Caregiver-centered perspective: the LLM adopts the viewpoint of the caregiver and reflects on their subjective experience. (2)Clinical psychometrician perspective: the LLM evaluates the caregiver’s risk level using an expert assessment lens.

We also implemented two task framings: (A) Direct classification: the LLM receives cutoff values and outputs a binary risk label, (B) Score prediction: the LLM predicts scale scores, which are then thresholded into binary labels. Under the caregiver perspective, the model self-assesses each scale item. Under the psychometrician perspective, the model estimates the total score directly.

Within the psychometrician perspective, we further compared: (a) Informed prompting: the full questionnaire is provided. (b) Uninformed prompting: only cutoff information is provided.

A summary of all prompting strategies is provided in S2 Table, and a schematic illustration is shown in S2 Fig. Zero-shot inference was conducted using Gemini 2.0, GPT-4o, and Llama 4.

### 2.7 Evaluation

The performance of traditional machine learning models and LLM-based approaches was evaluated using accuracy, recall, F1-score, and precision.

For LLM-based approaches, zero-shot predictions were directly compared with ground truth labels to compute evaluation metrics. To account for variability in LLM outputs, results are reported as the mean and standard deviation over five runs.

## 3 Results

### 3.1 Participant Characteristics

Participants (N = 32) were predominantly female (28/32), older adults (mean = 71, SD = 7.04) and retired (27/32). Descriptive statistics for the three psychological scales indicated moderate variability in perceived stress (PSS: mean = 16.78, SD = 6.28; range = 6–30), caregiver burden (ZBI: mean = 21.72, SD = 7.78; range = 8–37), and loneliness (UCLALS: mean = 41.38, SD = 9.83). Pairwise Pearson correlations (Fig. 2) showed moderate associations among the three constructs, indicating related but distinct dimensions of caregiver well-being. Across all scales, the majority of participants fell into the high-risk category (Table 1).

**Table 1.** Binary classification rules and label distribution for psychological scales.

| Scale | Scale Range | Binary Rule | Low Risk (n,%) | High Risk (n,%) |
| --- | --- | --- | --- | --- |
| PSS [10] | 0–40 | $\geq 14 \rightarrow 1$ | 10 (31.25%) | 22 (68.75%) |
| ZBI [11] | 0–52 | $\geq 21 \rightarrow 1$ | 12 (37.50%) | 20 (62.50%) |
| UCLALS [12] | 20–80 | $\geq 35 \rightarrow 1$ | 8 (25.00%) | 24 (75.00%) |

**Fig 2.**
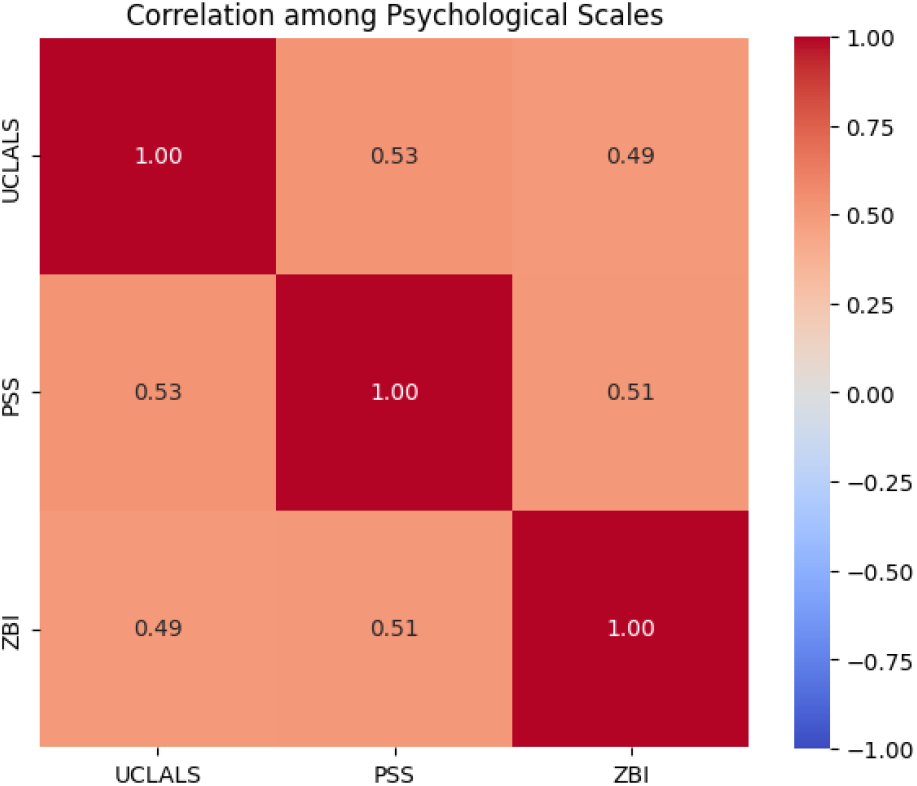
Pairwise correlations among PSS, ZBI, and UCLALS scores. The results show moderate correlations, suggesting that the three constructs are related but distinct.

### 3.2 Research Questions

We address the following research questions.

Q1. Which model (traditional machine learning vs. LLMs) performs best in detecting high and low PSS, ZBI, and UCLALS groups?

Q2. Which modality (wearable, interview, or their combination) performs best?

Q3. Which features are most informative for predicting each psychological outcome?

Q4. Which prompt engineering strategy (among the six proposed strategies) yields the best performance?

Q5. Which LLM (Gemini 2.0, Llama 4, or GPT-4o) achieves the best performance?

### 3.3 Overall Model and Modality Comparison (Q1 and Q2)

Across all three psychological scales, model performance varied by both model family and input modality. LLMs performed best when interview text was available, indicating that narrative responses contain emotional and contextual cues that these models can effectively use. In contrast, traditional machine learning models performed best in the multimodal setting, where structured wearable features complemented linguistic information. Wearable-only input produced the weakest performance for both model families.

PSS was the most predictable construct overall, with both ML and LLM approaches exceeding baseline performance. ZBI and UCLALS were more challenging, though LLMs showed relatively stronger performance on ZBI compared with traditional models. Representative PSS results are shown in Table 2, with full results for all scales provided in Supplementary Tables S3 Table, and S4 Table.

**Table 2.**
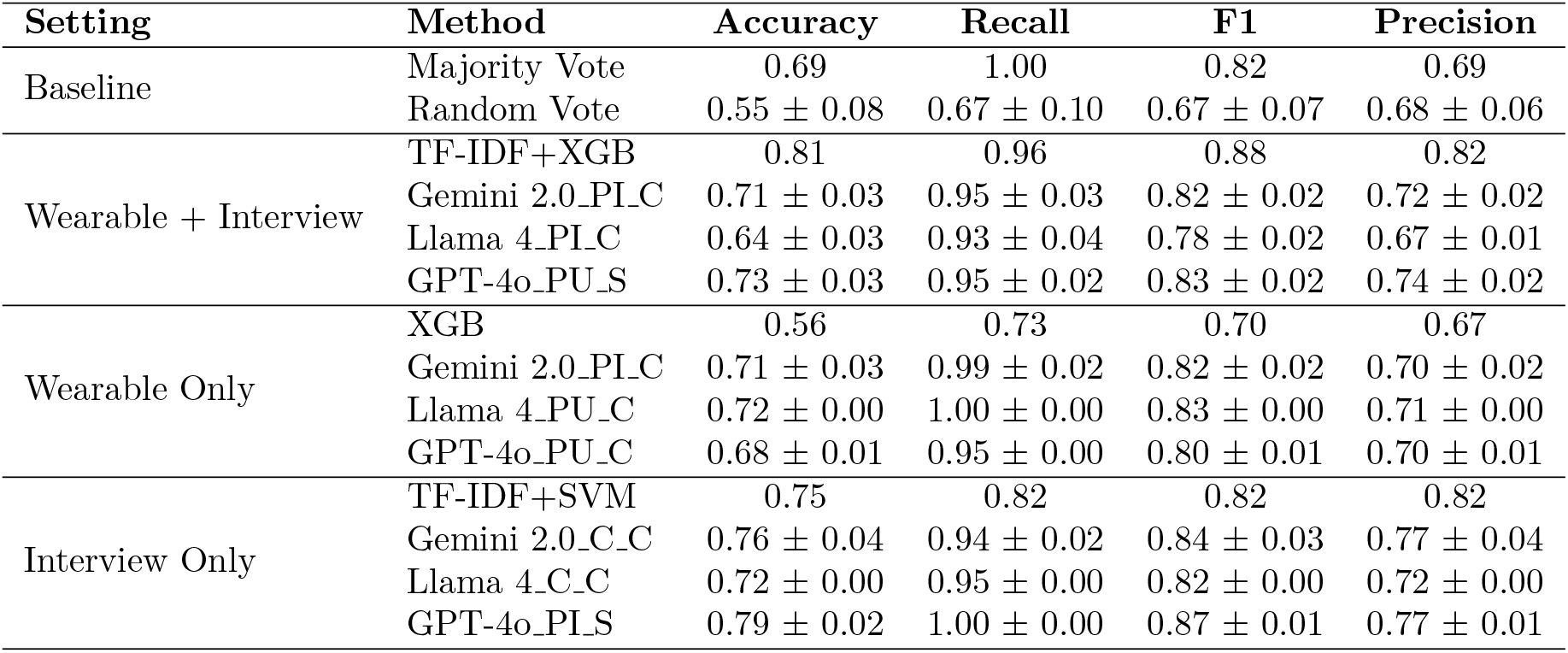
Performance comparison for PSS classification under different settings.

Table 3 shows the best performance across different modality and traditional machine learning configurations. Overall, the multimodal setting achieved the best performance and most consistent results across all three scales. For PSS and UCLALS, the multimodal configuration achieved the better performance (accuracy: 0.81 and 0.75, respectively), while the Interview-Only setting consistently outperformed the Wearable-Only setting. In contrast, for ZBI, Wearable-Only and Interview-Only configurations showed comparable performance, with only modest gains from multimodal integration. See S3 Fig for the comparison of machine learning models with the multimodal setting.

**Table 3.**
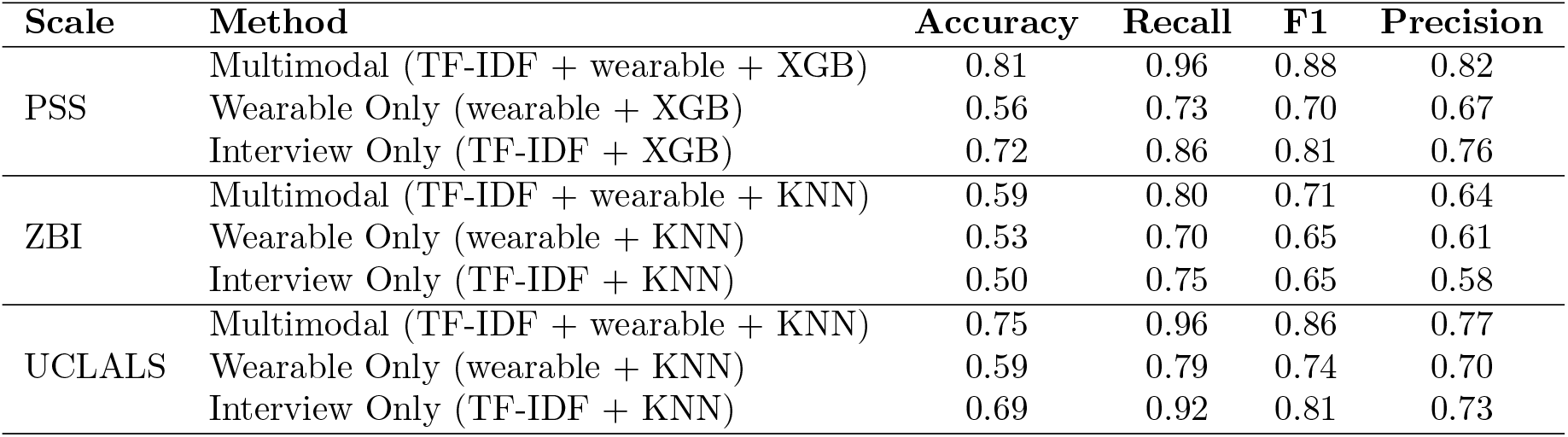
Performance of Traditional Machine Learning Methods Across Different Modalities.

| Scale | Method | Accuracy | Recall | F1 | Precision |
| --- | --- | --- | --- | --- | --- |
| PSS | Multimodal (TF-IDF + wearable + XGB) | 0.81 | 0.96 | 0.88 | 0.82 |
|  | Wearable Only (wearable + XGB) | 0.56 | 0.73 | 0.70 | 0.67 |
|  | Interview Only (TF-IDF + XGB) | 0.72 | 0.86 | 0.81 | 0.76 |
| ZBI | Multimodal (TF-IDF + wearable + KNN) | 0.59 | 0.80 | 0.71 | 0.64 |
|  | Wearable Only (wearable + KNN) | 0.53 | 0.70 | 0.65 | 0.61 |
|  | Interview Only (TF-IDF + KNN) | 0.50 | 0.75 | 0.65 | 0.58 |
| UCLALS | Multimodal (TF-IDF + wearable + KNN) | 0.75 | 0.96 | 0.86 | 0.77 |
|  | Wearable Only (wearable + KNN) | 0.59 | 0.79 | 0.74 | 0.70 |
|  | Interview Only (TF-IDF + KNN) | 0.69 | 0.92 | 0.81 | 0.73 |

One-way ANOVA and Tukey HSD tests showed significant modality effects in LLMs. (Fig. 3). We found LLM performed best with multimodal and interview-based inputs, and worst with Wearable-Only inputs, particularly for UCLALS and ZBI.

**Fig 3.**
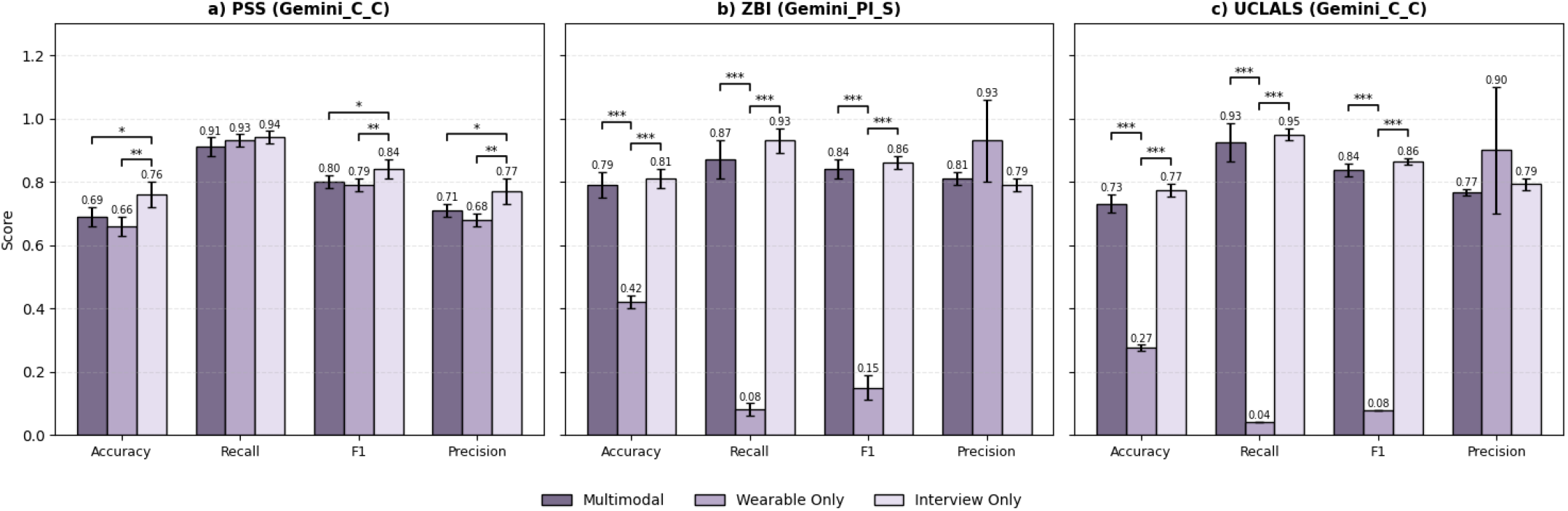
Performance comparison across three modalities (Gemini 2.0); significance tested via ANOVA and Tukey HSD.

We also compared the impact of input representation by providing LLMs with either raw interview transcripts or the top-20 TF-IDF features. Raw Interview input consistently outperformed TF-IDF representations across all prompting strategies. Under the multimodal setting, improvements are moderate (approximately +0.03 in accuracy and +0.02–0.03 in F1), whereas under the Interview-Only setting, gains are more substantial (approximately +0.06 in accuracy and +0.03–0.04 in F1). This suggests that preserving contextual and narrative information is particularly important for LLM-based reasoning.

### 3.4 Feature Contributions in Traditional Machine Learning (Q3)

The above analysis shows that traditional machine learning models achieved the best performance under the multimodal setting. To further understand which features contributed most to model predictions, we conducted a SHAP-based feature importance analysis using XGBoost under the Multimodal setting for all three scales .

As shown in Fig. 4, PSS predictions were mainly driven by TF-IDF linguistic features related to descriptions of daily routines and time-related experiences, such as “time” and “long”, along with wearable activity-related measures including average daily maximum minute-level step count (*steps_max*) and activity levels during the least active 5-hour period (*Steps_L5*). The high PSS group showed lower minute-level maximum step counts compared to the low PSS group.

**Fig 4.**
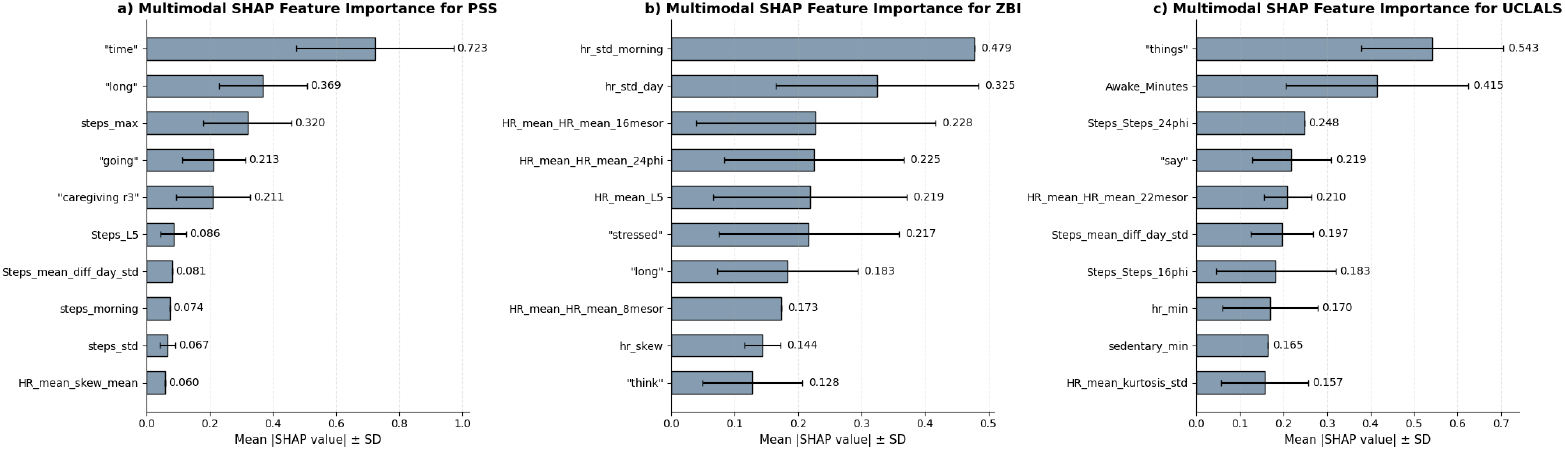
SHAP-based feature importance for the multimodal setting across PSS, ZBI, and UCLALS. Values represent mean absolute SHAP values aggregated over LOOCV folds using multimodal features derived from wearable and interview data.

ZBI predictions were primarily associated with heart rate variability and rhythm features, including morning heart rate variability (*hr_std_morning*), afternoon heart rate variability (*hr_std_afternoon*), and the 16-hour heart rate mesor. Linguistic features related to psychosocial strain, such as “stressed” and “think”, also showed relatively high importance.

In contrast, UCLALS predictions were mainly driven by sleep disturbance and activity-related rhythm features, with high UCLALS group showing higher awake minutes (*Awake_Minutes*) and delayed circadian step rhythm patterns compared to the low UCLALS group. Several linguistic features related to communication and everyday expression, including “things” and “say”, also contributed to prediction performance.

### 3.5 Effect of Prompt Engineering Strategies (Q4)

Due to the difficulty of identifying a consistently superior prompt engineering strategy across all settings, we further conducted a focused analysis of two representative scenarios: ZBI (multimodal), where LLMs showed relatively better performance (S5 Table), and UCLALS (Wearable-Only), where LLM performance was comparatively lower ( S6 Table).

For ZBI under the multimodal setting, all prompt strategies achieve relatively strong performance, with the caregiver-centered classification approach (Gemini_C_C) attaining the highest accuracy (0.81). The preliminary ANOVA and Tukey results suggest differences across strategies in accuracy (*F* = 5.36, *p* = 0.0019), recall (*F* = 7.49, *p* < 0.001), and precision (*F* = 5.56, *p* = 0.0015) with PI S showing more consistent performance and PU C performing worse overall.

For UCLALS under the Wearable-Only setting, only Gemini_C_S (caregiver-centered score-based prompting) achieved reasonable performance. Other strategies performed considerably worse. To better understand this discrepancy, we analyzed the reasoning outputs of the best-performing model (Gemini_C_S) and the worst-performing model (Gemini_PI_S) using a simple lexical feature analysis capturing hedging, intensity descriptors, and negation. As shown in S4 Fig, the better-performing model more frequently used moderate-level descriptors (0.84 vs. 0.57) and hedging language (0.42 vs. 0.27), suggesting that more uncertainty-aware reasoning may better align with the subjective nature of loneliness assessment.

### 3.6 Comparison across LLMs (Q5)

To enable a fair comparison across LLMs, we evaluated Gemini 2.0, GPT-4o, and Llama 4 under the Interview-Only setting, using the same prompting strategy for each scale (S5 Fig). Overall, Llama 4 showed slightly lower performance than Gemini 2.0 and GPT-4o on PSS and ZBI, while achieving comparable performance on UCLALS, possibly due to class imbalance in the UCLALS dataset. GPT-4o achieved the best performance on PSS (accuracy: 0.79, F1: 0.87), whereas Gemini 2.0 performed best on ZBI and UCLALS, including the highest accuracy on ZBI (0.81). Similar patterns were observed across Tables 2, S3 Table, and S4 Table, with relative LLM performance remaining generally consistent across prompting strategies and modality settings.

## 4 Discussion

This study provides a preliminary systematic comparison of traditional machine learning and LLM-based approaches for identifying risk groups among AD/ADRD caregivers across PSS, ZBI, and UCLALS and multiple data modalities. The results highlighed several meaningful findings.

First, PSS was consistently more predictable construct than ZBI and UCLALS across model families and modalities. This predictability suggests that the PSS captures a generalized latent construct of distress that manifests reliably across disparate behavioral and physiological channels. Unlike scales designed to capture specific syndromic domains (such as social isolation or caregiving burden), the PSS measures a global appraisal of uncontrollability and overload [10]. Because this cognitive appraisal fundamentally alters both top-down behavioral regulation and bottom-up autonomic output, its signatures are embedded within multi-modal data streams such as language and behavior, facilitating machine learning classification performance.

Our feature analysis also showed differences in how three constructs manifest in behavior and language. For PSS, words like “time” and “long” can serve as proxies for acute time pressure and emotional overload and reflect scenarios where environmental demands exceed personal resources [53], a core tenet of transactional stress theory that links high cognitive load to negative stress appraisal. Reduced activity peaks and diurnal rhythms (e.g., lower maximum minute-level step counts and reduced activity during the least active 5-hour period (L5)) can reflect chronic stress, psychological fatigue, and a hyperactive sympathetic nervous system that prevents natural recovery cycles as reported in prior work [54, 55].

In contrast, for ZBI, HRV and macro-level rhythm features are informative features. This can align with psychisomatic evidence that caregiver burden is tied to long-term autonomic disregulation [56]. Linguistic indicators of psychological strain (e.g., “stressed”) can further suggest increased rumination and emotional overload related to caregiver burden. UCLALS reflects social-emotional isolation and can affect sleep disturbance and physical activity rhythms and vague relational, less emotionally explicit language [57] .

Second, classification performance was shaped by data modality. Traditional machine learning models achieved the strongest performance under the multimodal setting, suggesting that combining wearable and interview data captured complementary physiological, behavioral, and linguistic signals. In contrast, LLMs achieved their strongest performance with Interview-Only data, highlighting their ability to extract nuanced emotional and contextual cues from narrative text. The relatively limited improvement of LLMs under multimodal settings suggests that simply combining wearable features with interview data may introduce redundant or noisy information that reduces the effectiveness of model reasoning.

Third, prompt engineering influenced LLM performance. Although no consistent pattern was found across different prompt engineering strategies, ANOVA results show that different prompting approaches lead to statistically significant differences across multiple evaluation metrics. This finding underscores the importance of prompt design as a critical component in optimizing LLM-based predictive systems.

Lastly, LLM performance varied across models. Gemini 2.0 showed the best and most stable performance. In contrast, GPT-4o consistently struggled under the Wearable Only setting, which may be due to its limited ability to effectively utilize structured numerical features. Similarly, Llama 4 also underperformed compared to Gemini 2.0 and GPT-4o across multiple tasks, possibly because it is less effective in both structured data processing and complex reasoning.

This study extends prior research on AI-based mental health assessment in several ways. Unlike most prior work that relies on either social media text [58, 59] or physiological signals [20, 21] alone, we evaluate Wearable-Only, Interview-Only, and Multimodal configurations using real-world data from AD/ADRD caregivers. Prior LLM studies have primarily focused on generic prompting strategies such as zero-shot or few-shot prompting [38, 39]; in contrast, we evaluate role-aware prompting (caregiver-centered vs. psychometrician-centered) and task framing (direct classification vs. score prediction), demonstrating that prompting perspective and framing can meaningfully influence model outputs. In addition, while many previous studies focus on a single psychological construct [20, 58], our study jointly evaluates PSS, ZBI, and UCLALS, enabling comparison of predictability across different dimensions of caregiver well-being. Finally, by comparing traditional machine learning and LLMs under matched experimental settings, our findings provide practical insights into how different model families perform across modalities and tasks.

This study has several limitations. First, the sample size is limited, which may affect the generalizability of the findings. Although LOOCV mitigates some concerns, larger datasets are needed to validate these findings. In addition to wearable data, other data sources such as mobile phone sensing data may provide richer behavioral and contextual information in future studies. Second, the multimodal setting used in this study was intentionally simple to maintain interpretability. More advanced multimodal fusion methods may better capture complementary information across wearable and interview data. Third, prompt engineering effects were inconsistent and difficult to fully interpret, reflecting the broader challenge that LLM behavior is often sensitive to subtle prompt design choices. Further research is therefore needed to develop more consistent and interpretable prompting strategies. Finally, the study focused on binary risk classification, which may oversimplify the nuanced experiences of caregivers. Future work could explore continuous score prediction or multi-level risk stratification.

## Data Availability

Anonymized data will be available upon request through Zenodo.

## Supporting information

**S1 Table.** Summary of top features across PSS, UCLALS, and ZBI scales. The first column indicates which scale(s) the feature is associated with, followed by the feature name and its description.

| Scale | Feature | Description |
| --- | --- | --- |
| PSS | steps_afternoon | Mean daily step count during the afternoon (12:00–17:59) |
| PSS | steps_max | Mean of the daily maximum step count recorded in one minute |
| PSS & UCLALS | Restless_Minutes | Mean nightly minutes of restlessness during sleep |
| PSS | hr_iqr | Mean daily interquartile range of minute-level heart rate |
| PSS | HR_mean_HR_mean_24phi | Acrophase of the 24-hour heart rate rhythm |
| UCLALS | Awake_Minutes | Mean nightly minutes awake during sleep |
| UCLALS | HR_mean_kurtosis_std | Cross-day variability of HR deviation kurtosis |
| UCLALS | Steps_sd_diff_day_std | Cross-day variability of Steps deviation SD |
| UCLALS | SRI_multistage | Multistage sleep regularity index |
| ZBI | HR_mean_L5 | Average heart rate during the lowest 5-hour night period |
| ZBI | HR_mean_HR_mean_16mesor | Baseline heart rate from a 16-hour cosinor model |
| ZBI | HR_mean_HR_mean_20mesor | Baseline heart rate from a 20-hour cosinor model |
| ZBI | HR_mean_HR_mean_22mesor | Baseline heart rate from a 22-hour cosinor model |
| ZBI | HR_mean_HR_mean_24mesor | Baseline heart rate from a 24-hour circadian cosinor model |

**S2 Table.** Summary and abbreviations of the six prompt engineering strategies used in this study.

| Strategy | Description |
| --- | --- |
| Caregiver Score Prediction (C_S) | Caregiver perspective; predicts a questionnaire score and converts it to a binary risk label using the corresponding cut-off. |
| Caregiver Direct Classification (C_C) | Caregiver perspective; directly predicts a binary risk label. |
| Psychometrician Informed Score Prediction (PI_S) | Psychometrician perspective with questionnaire information; predicts a questionnaire score and converts it to a binary risk label using the corresponding cut-off. |
| Psychometrician Informed Classification (PI_C) | Psychometrician perspective with questionnaire information; directly predicts a binary risk label. |
| Psychometrician Uninformed Score Prediction (PU_S) | Psychometrician perspective without questionnaire information; predicts a questionnaire score and converts it to a binary risk label using the corresponding cut-off. |
| Psychometrician Uninformed Classification (PU_C) | Psychometrician perspective without questionnaire information; directly predicts a binary risk label. |

**S3 Table.**
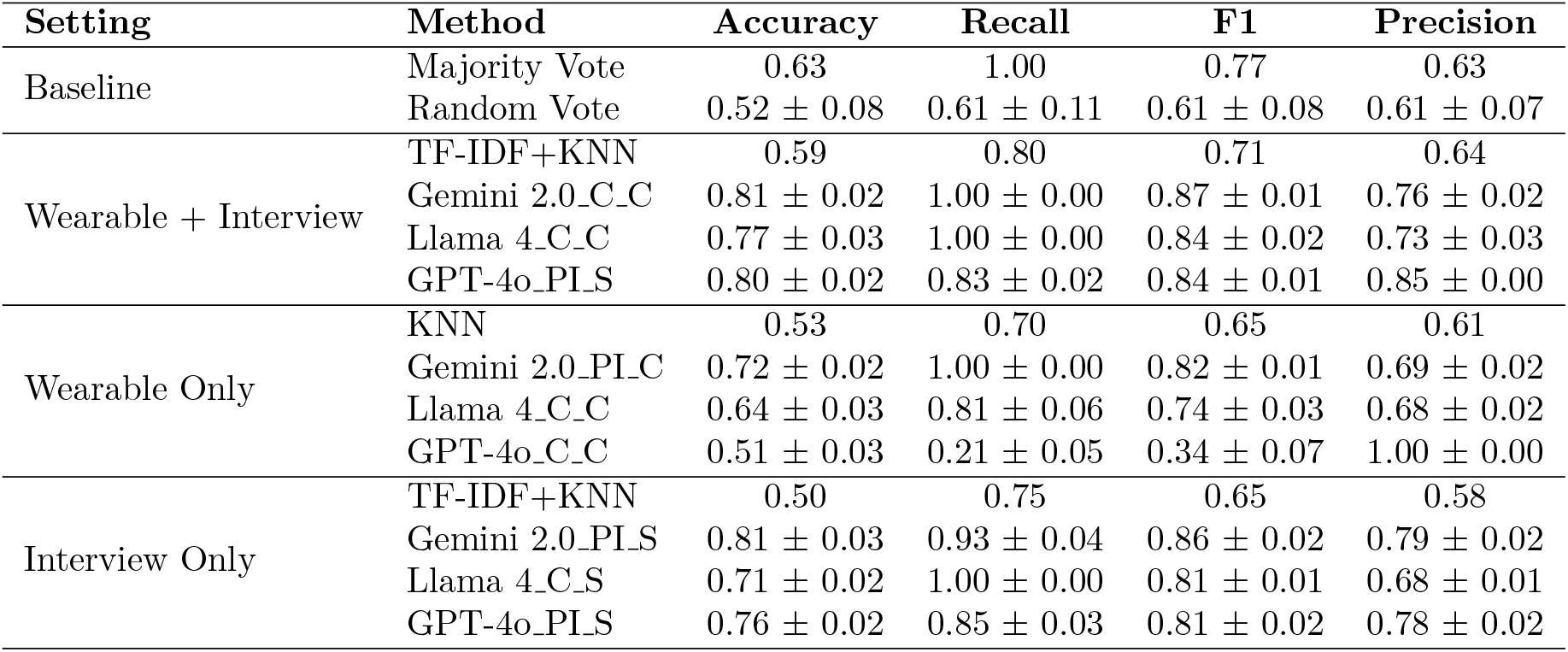
Performance comparison for ZBI classification under different settings. The table reports accuracy, recall, F1 score, and precision across different input modalities and modeling approaches.

**S4 Table.**
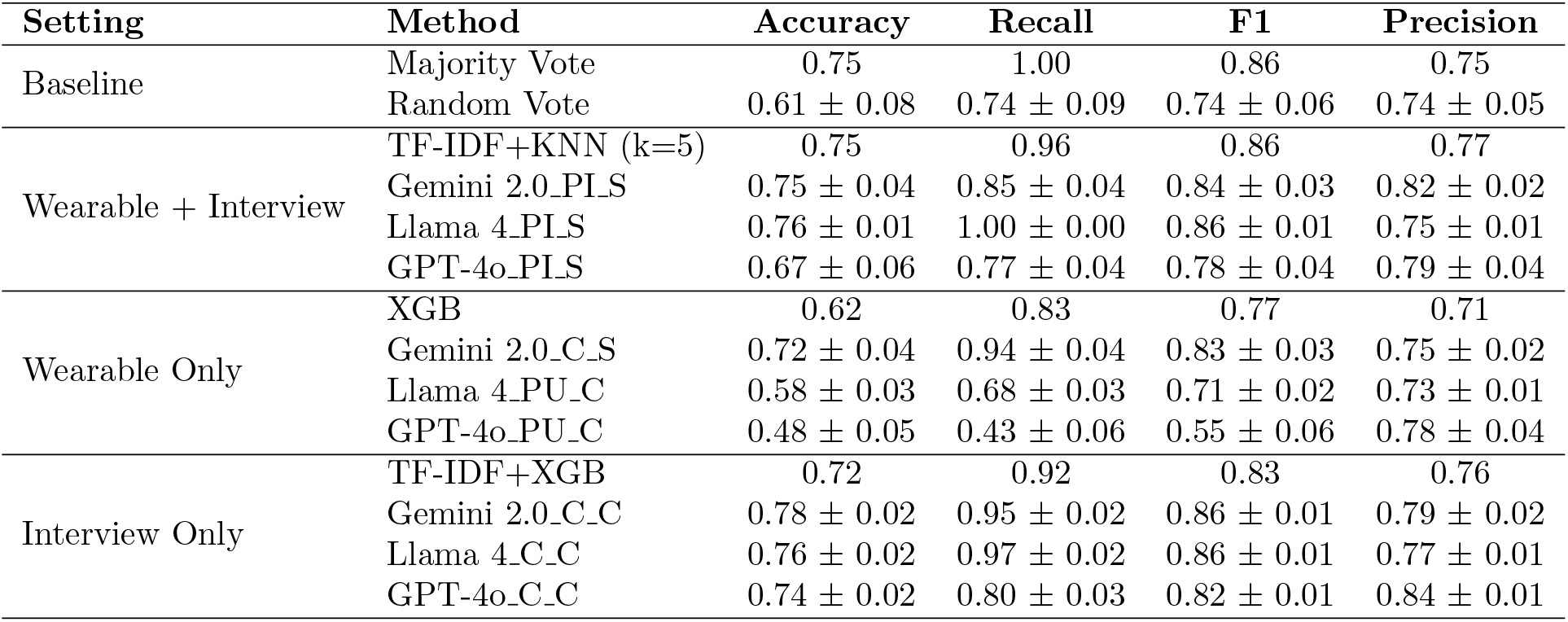
Performance comparison for UCLALS classification under different settings. The table reports accuracy, recall, F1 score, and precision across different input modalities and modeling approaches.

**S5 Table.**
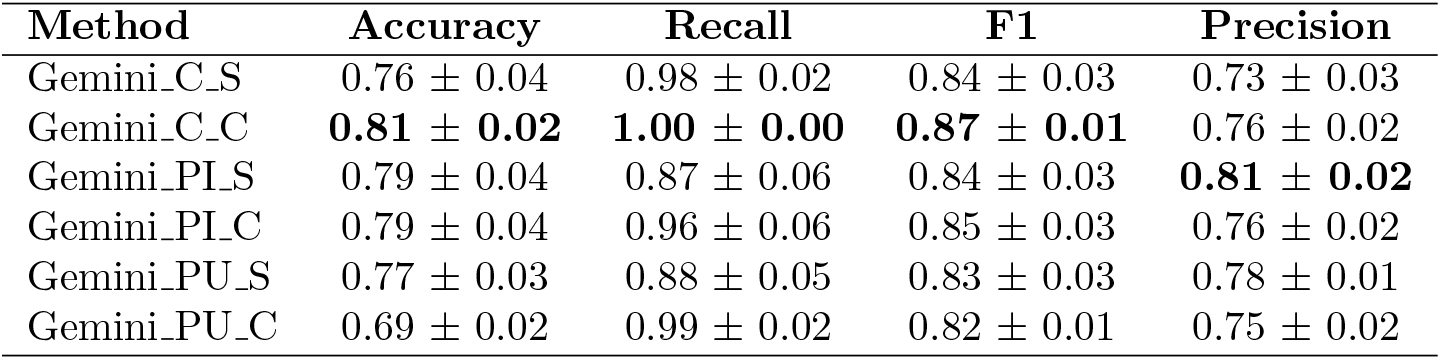
Performance comparison of different prompt engineering strategies for ZBI under multimodal setting.

**S6 Table.**
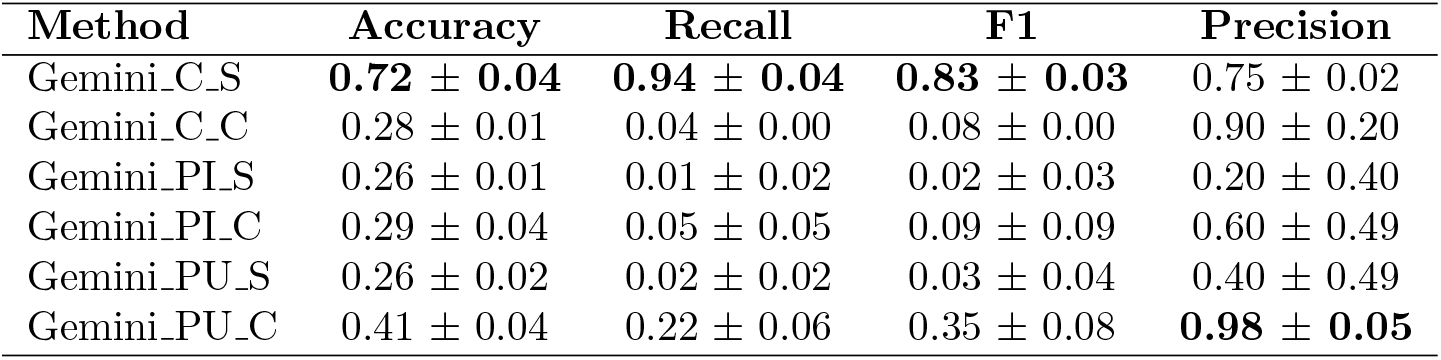
Performance comparison of different prompt engineering strategies for UCLALS under Wearable-Only setting.

**S1 Fig.**
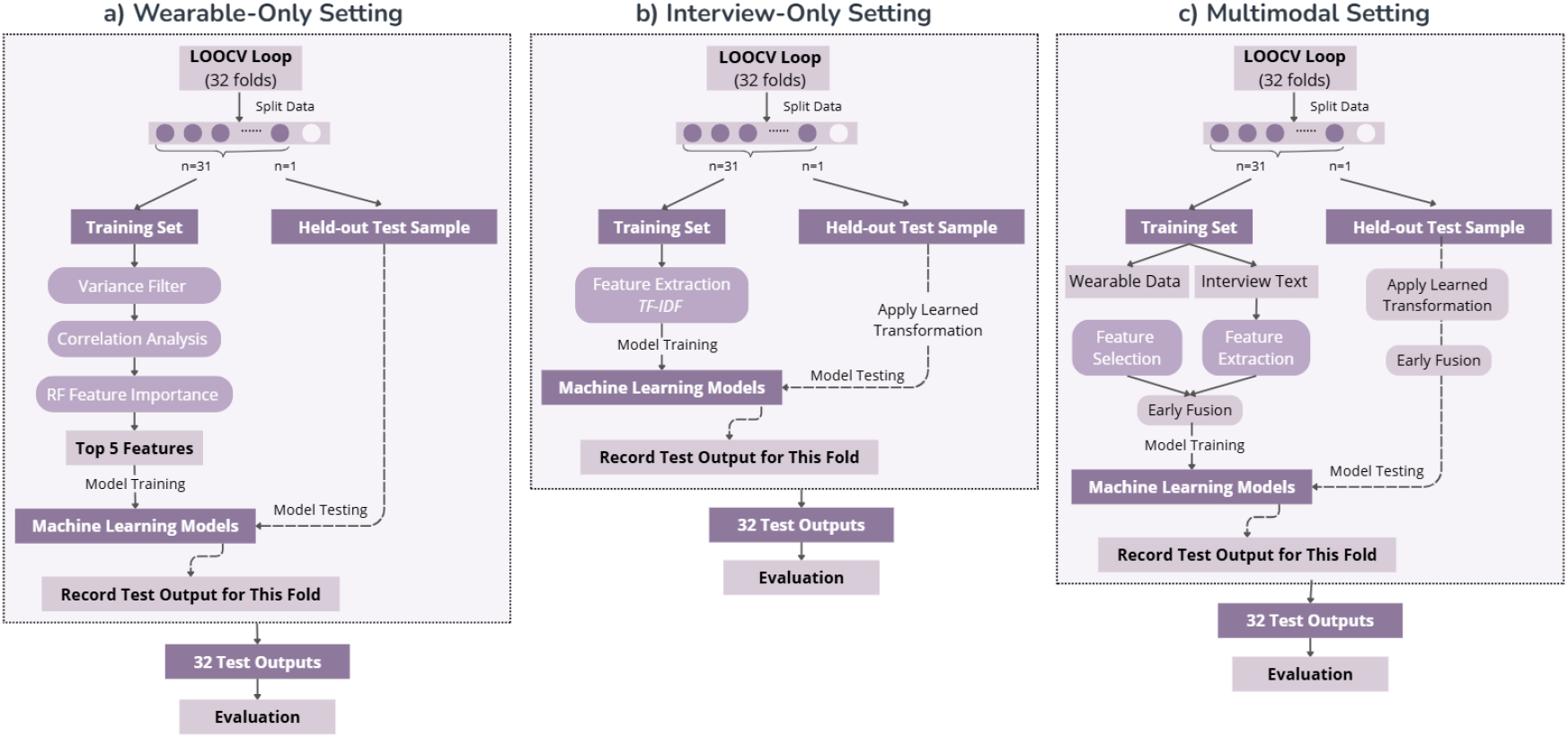
LOOCV-based machine learning models training and evaluation pipeline.

**S2 Fig.**
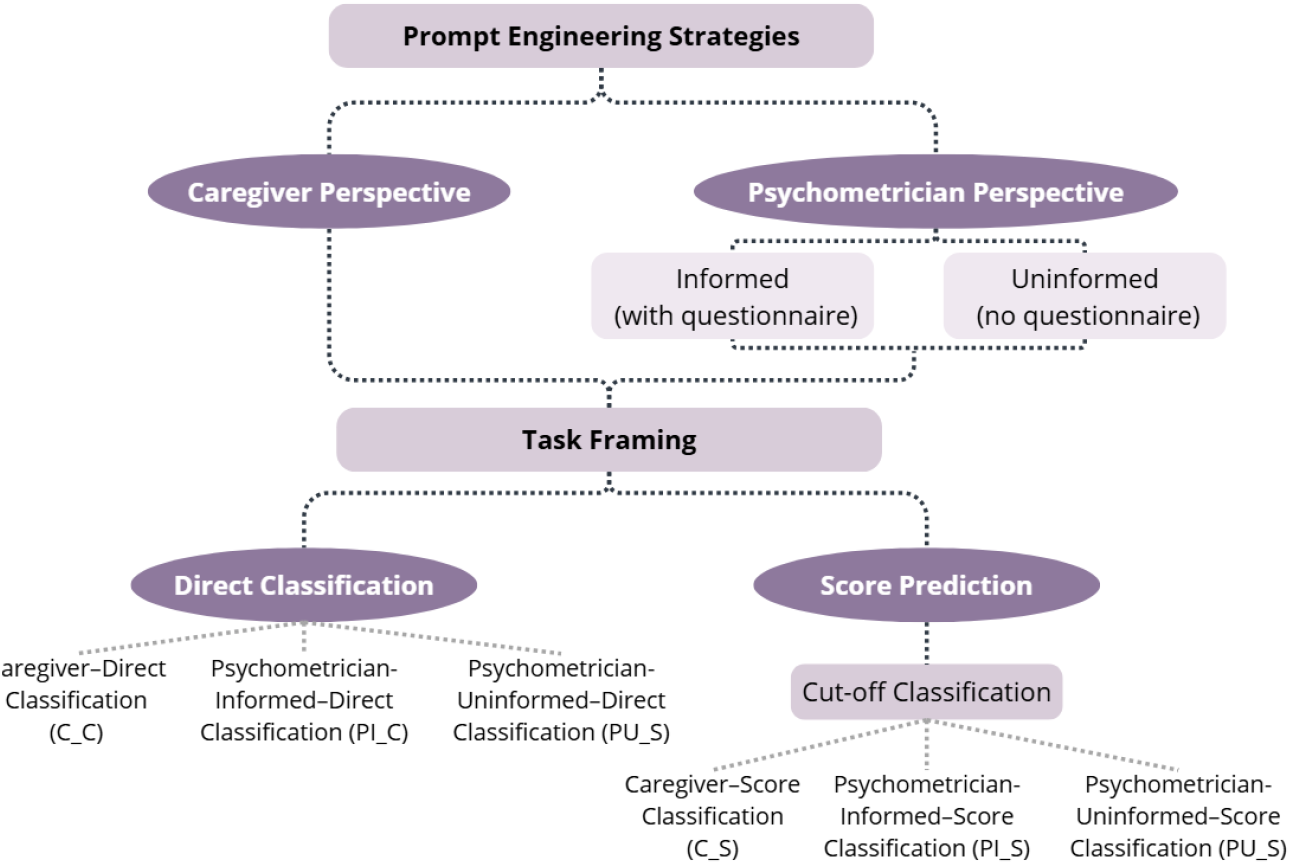
Schematic illustration of different prompt engineering strategies, including variations in perspective and access to questionnaire information.

**S3 Fig.**
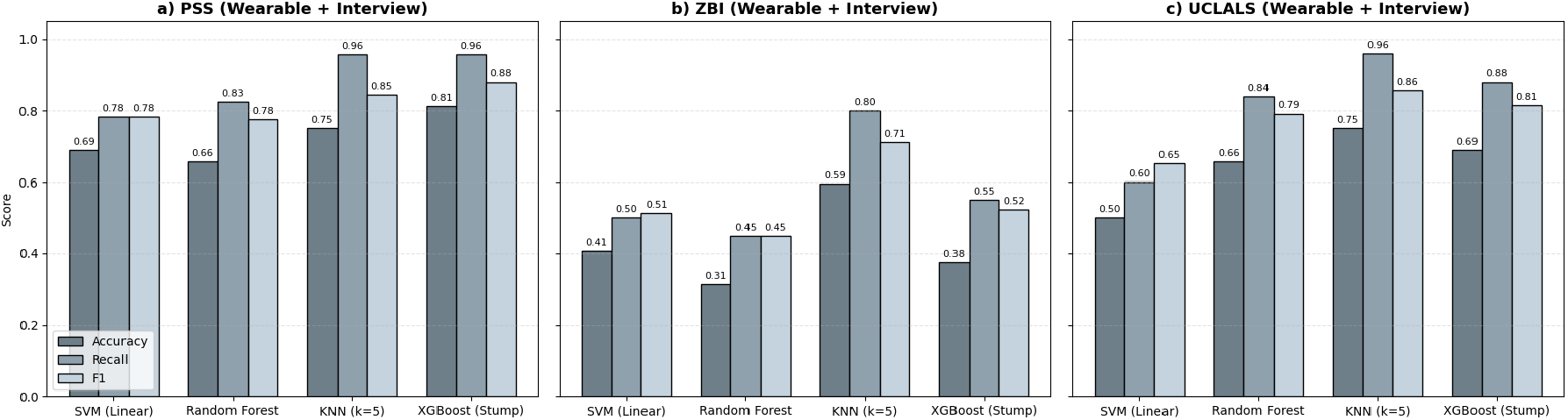
Comparison of machine learning models across PSS, ZBI, and UCLALS under the multimodal setting.

**S4 Fig.**
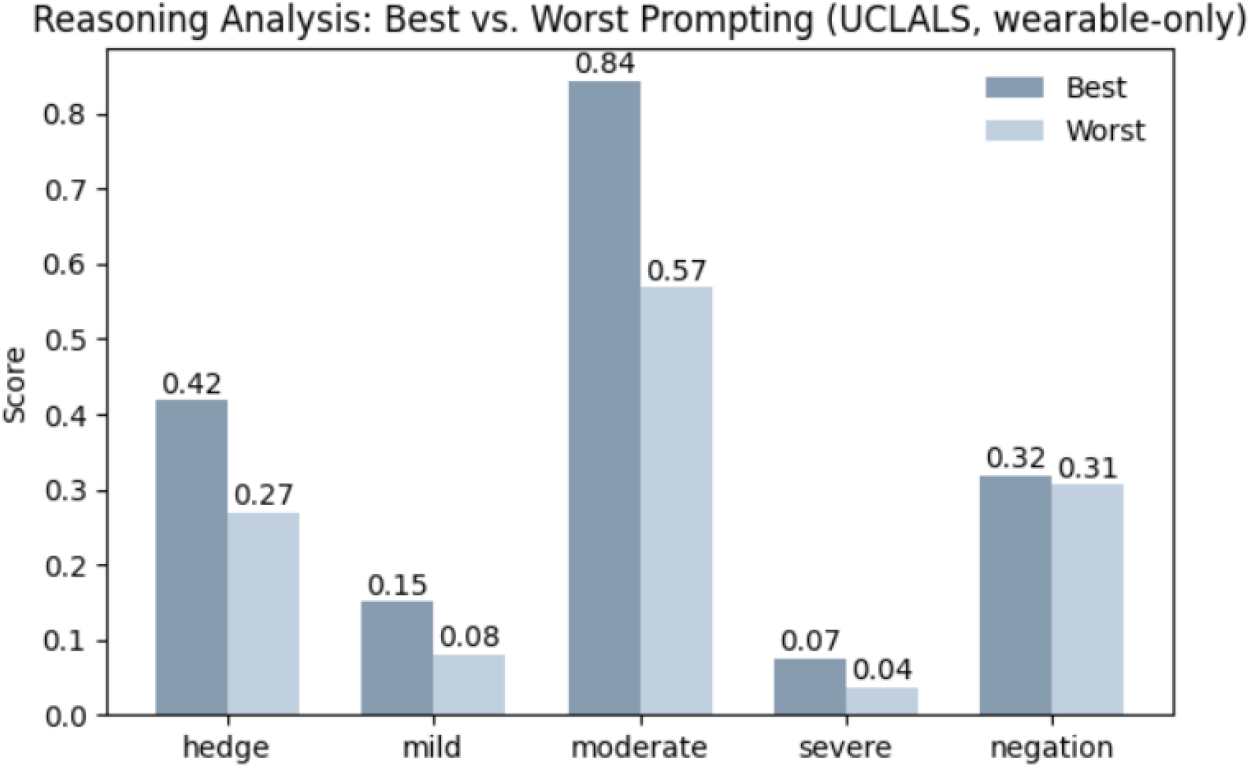
Comparison of lexical features in reasoning outputs between the best-performing (Gemini C S) and worst-performing (Gemini PI S) models.

**S5 Fig.**
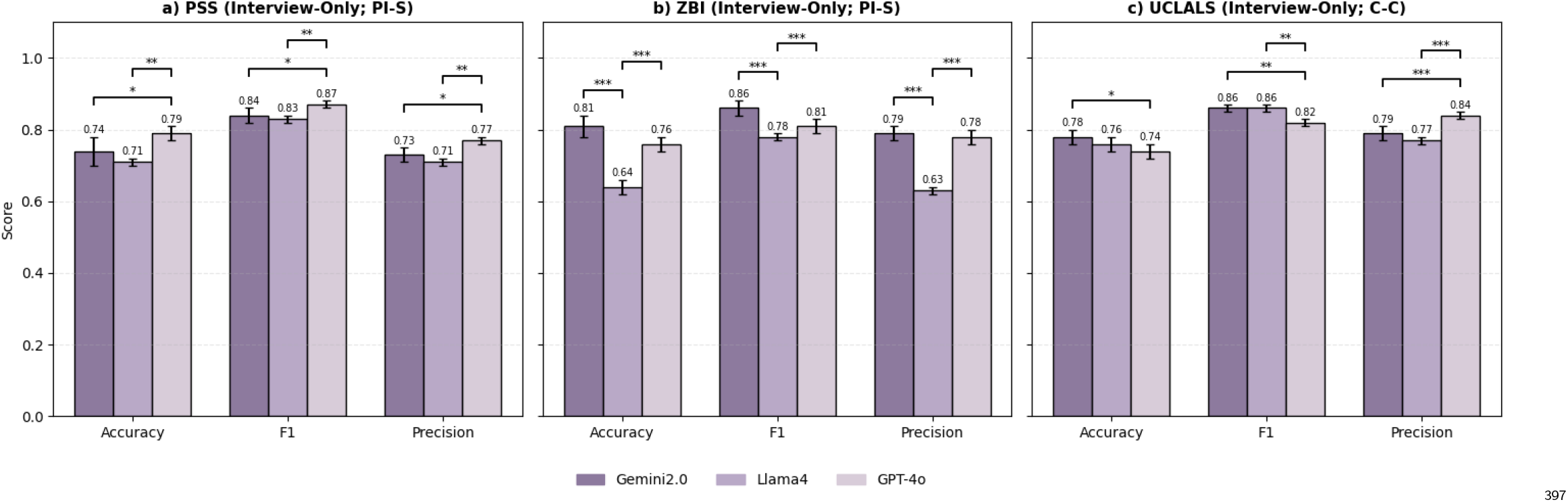
Comparison of LLM performance across PSS, ZBI, and UCLALS under the Interview Only setting.

## Notes

### Competing Interest Statement

The authors have declared no competing interest.

### Funding Statement

Yes

### Author Declarations

The study is approved under Rice University Institutional Review Board (IRB).

